# Using Machine Learning to Classify Schizophrenia Based on Retinal Images

**DOI:** 10.1101/2021.04.04.21254893

**Authors:** Diana Joseph, Adriann Lai, Steven Silverstein, Rajeev Ramchandran, Edgar A. Bernal

## Abstract

VII.

**Objectives:** Thinning of retinal layers has been documented in patients with chronic schizophrenia using standard metrics of optical coherence tomography (OCT) devices. We demonstrate the effectiveness of machine learning (ML) techniques to differentiate between schizophrenia patients and healthy controls using OCT images.

**Methods:** Features extracted from a convolutional neural network (CNN) designed to segment retinal layers from OCT images represented abstracted data from the OCT images of 14 first episode (FEP) and 18 chronic schizophrenia patients, and their respective 20 and 18 age-matched controls. The abstracted data and OCT machine metrics were used separately to train support vector classification (SVC) models to differentiate between control and schizophrenia samples and test them.

**Results:** SVCs operating on OCT machine metrics did not classify unseen samples of FEP schizophrenia patients and controls with performance better than chance, while those looking at chronic schizophrenia did, paralleling results obtained using parametric statistics. In contrast, SVCs operating on OCT image data extracted from the CNN classified unseen samples from both populations with performance greater than chance.

**Conclusion:** These results suggest that ML techniques can detect patterns in patients with FEP schizophrenia with greater performance using features extracted from OCT images than metrics provided by OCT machines.

## VIII. Introduction

Machine Learning (ML) techniques enable computers to learn from data. Most ML applications fall under the category of supervised learning, whereby an algorithm is trained with pairs of data samples and corresponding labels and learns to assign labels to unseen samples (Bishop 2006). Another area where ML algorithms are effective is in the discovery, in a bottom-up or data-driven fashion, of novel relationships in multidimensional data (Bishop 2006). This approach has demonstrated effectiveness in multiple fields (Bishop 2006, LeCun et al., 2015) ranging across financial services (e.g., stock market analysis (Lopez de Prado, 2018), fraud detection (Chalapathy et al., 2019)), clinical applications (e.g., disease diagnosis (Heinrichs et al., 2019) and progression prediction (Wang, 2019)), drug discovery (Ekins et al., 2019), robotics (Wang et al., 2019, Oshin et al., 2019), and social media (e.g., social network data analysis (Jaffali et al., 2019) and content filtering (Dada et al., 2019)). Within the broader healthcare field, one area where ML may be useful is in the discovery of biomarkers (e.g., predictors of illness development or course, see Leclerq et al., 2019). In this study, we focus on the exploration of novel biomarkers in neuropsychiatry. In the fields of human clinical neuropsychology, psychophysiology, and cognitive neuroscience, performance on experimental measures is typically measured in terms of variables such as accuracy, reaction time, waveform amplitude, or change in the blood oxygen-level dependent (BOLD) signal. Similarly, in studies of central nervous system (CNS) structure, typical parameters involve 2D thickness values, or 3D volumes. With the advent of ML-based approaches, however, comparisons between groups (e.g., a patient group and a control group) no longer need to be limited to those involving traditional metrics. With ML, it is possible to search for biomarkers based on complex patterns of relationships between multiple variables, including patterns that had not been previously considered. In traditional ML literature, patterns driven by heuristics or domain knowledge are termed ‘hand-crafted’ or ‘hand-engineered’, in contrast with ‘data-driven’ or ‘learned’ representations. It is widely accepted by the ML community that the latter tend to be most effective at automating decision-making and discovery, particularly in scenarios where abundant and complex data are available (LeCun et al., 2015, Goodfellow, 2016).

In this study, we sought to demonstrate the potential effectiveness of an ML-based approach to investigating changes in retinal structure, as observed using optical coherence tomography (OCT; a form of retinal imaging) in people with schizophrenia. We undertook these analyses based on multiple considerations, including: 1) the retina is part of the CNS that develops out of the same tissue as the brain and shares many features (e.g., neurons and neurotransmitters, a layered architecture); 2) thinning of retinal neural and vascular layers has been identified in a range of neuropsychiatric disorders (e.g., Alzheimer’s disease, Parkinson’s disease, multiple sclerosis, traumatic brain injury) where it is correlated with, and often precedes, features such as brain volume loss, cognitive decline, and overall illness progression (reviewed in Silverstein et al., 2020a); 3) retinal thinning has been consistently observed in schizophrenia (reviewed in Silverstein et al., 2020b); but 4) findings regarding which layers demonstrate thinning have been inconsistent; and 5) partly as a result of this inconsistency, the overall effect sizes from between-group comparisons (e.g., schizophrenia vs. healthy control) are small or very small for all variables that have been measured to date (Kazakos & Karageorgiou, 2020; Silverstein et al., 2020b). This situation suggests two potential hypotheses and directions. One is that while thinning of retinal layers is reliably observed in schizophrenia, the specific layer(s) that are affected may be significantly determined by stochastic factors that are independent of the illness itself, as is thought to occur regarding the specific site of epileptic foci (Silverstein et al., 2020a). A second is that abnormalities in retinal structure in schizophrenia may be better characterized by variables other than overall thickness or volume. For example, it may be that there are changes to the internal structure of retinal layers that are primary, and that often, but not always, lead to overall thinning. Because it is not yet clear what such changes might be, however, a bottom-up approach that searches for patterns within the data is called for.

In this study, we report on the use of such an ML-based approach to differentiate schizophrenia patients from healthy controls. Specifically, we used data from a recently published study (Lai et al., 2020) in which (as in prior studies) chronic schizophrenia patients could be discriminated from age-matched controls on standard OCT metrics, but first episode schizophrenia patients (who had not been previously studied) could not be differentiated from their age-matched controls based on the same metrics. The data from Lai et al. was interpreted as evidence that retinal thinning and volume loss in schizophrenia are aspects of illness progression, and that they are not present at the initial episode of psychosis. However, we recognized the possibility that examination of patterns within the extensive image data generated by OCT might yield additional insights regarding group differences. Therefore, the purpose of this study was to determine the classification accuracy of ML methods and to examine this accuracy relative to results obtained from traditional OCT metrics analyzed with traditional parametric statistics to determine between group differences.

## Method

### Data Collection

Our data was collected as part of a previously published OCT study that examined and compared retinal structural differences between chronic and first episode schizophrenia spectrum disorder patients (Lai et al., 2020). The subjects included 14 first episode (FEP) schizophrenia patients and 18 chronic schizophrenia patients. There were also 20 age-matched controls for the FEP group and 18 age-matched controls for the chronically ill group. For each subject, we obtained data, from both eyes, on macula thickness and volume, combined ganglion and inner plexiform layer thickness measured at the macula, and retinal nerve fiber layer thickness measures adjacent to the optic nerve head. Data was acquired using a Spectral Domain Cirrus 5000 HD-OCT scanner. For this study, the data used included the values provided by the Cirrus machine for the above variables, and the data inherent to the images produced by the device.

### Strategy for Utilizing Neural Networks

A convolutional neural network (CNN) is a type of deep learning architecture that is often used for automated analysis of image data. Examples of tasks commonly carried out by CNNs include object detection, image segmentation and image classification. Because our study looked at OCT images, we decided to use a CNN. However, training a CNN requires a large data set, and our data was limited to images from each eye of the 70 subjects. Therefore, we relied on transfer learning techniques. Specifically, we leveraged the publicly available PyTorch implementation of ReLayNet, a pre-trained CNN that was designed to segment the macula layers of and identify pathologic fluid build up in OCT images from people with diabetic macular edema (Roy et al., 2017). The ReLayNet algorithm is an instance of a U-Net (Ronneberger et al., 2015), which is a type of CNN that involves encoding and subsequently decoding image data to produce multiple intermediate image feature convolutional layers. Each convolutional layer involves higher levels of abstraction. ReLayNet has seven convolutional layers between the OCT image input and the segmented output, and we looked at the middle three layers (*e*3, *bn*, and *d*3) for our analysis (see Figure 1). We chose these layers because we wanted intermediate activations that had been moderately processed but were not overly tuned to the segmentation task. This is because our goal was not to segment the OCT images, but rather use abstracted intermediates to classify the images as belonging to “control” or “schizophrenia” subjects.

**Figure 1.**
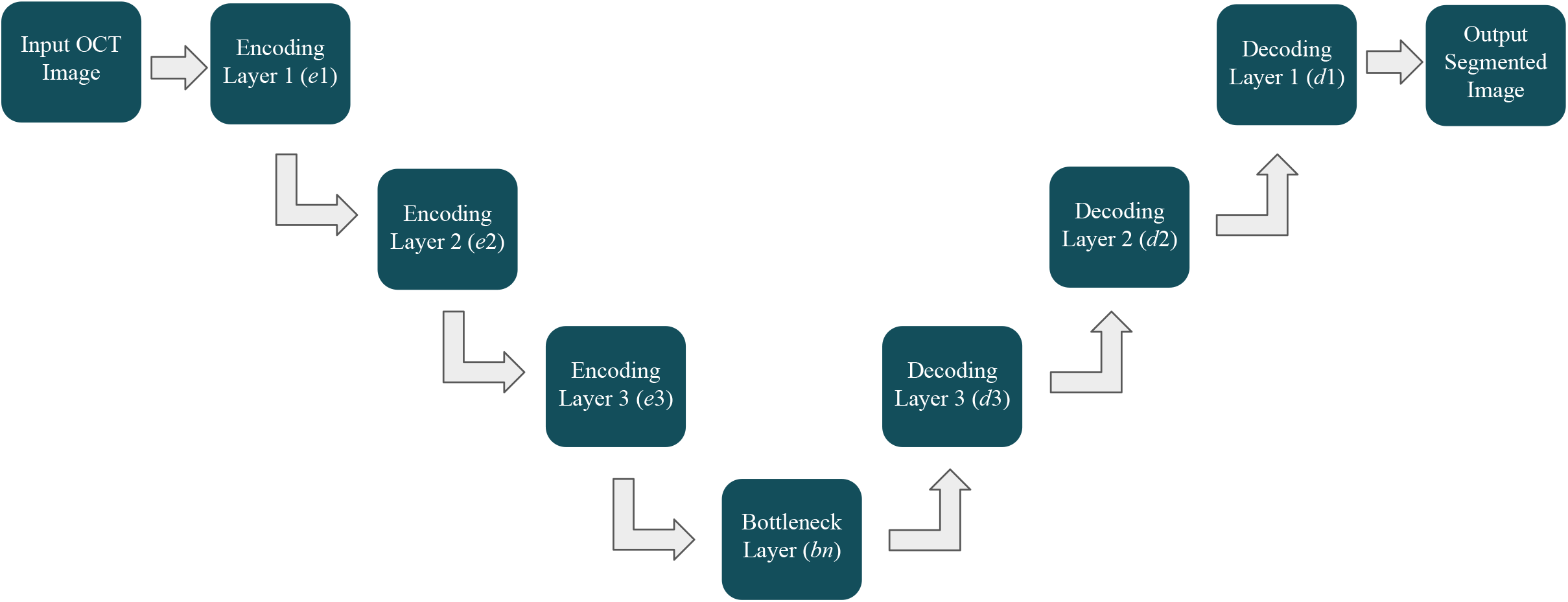
Outline of ReLayNet U-Net algorithm architecture. Each arrow represents a layer of encoding or decoding that furthers the abstraction of the convolutional layer from the input OCT image. The diagram illustrates the layer labeling convention used throughout the paper.

### Convolutional Layer Extraction

In order to address domain gaps that may exist between the imagery with which ReLayNet was trained and our own image data, we preprocessed our OCT images before feeding them to the pre-trained network. The first preprocessing step was to adjust the exposure of our OCT images so that they matched the exposure of the training images. We did this in Python 3.8 using the publicly available scikit-image project (Walt et al., 2014). We specifically used the `match_histograms` function in the exposure module. We then denoised the images using BM3D 3.0.6 software available for scientific research, with a noise standard deviation parameter of 30/255 and hard thresholding (Makinen et al., 2020). Lastly, our images were 508×338 pixels while the training set images were 512×496 pixels, and so we rescaled our images to have a height of 496 using MatLab’s `imresize` function and then evenly cropped each side to create a width of 512.

After preprocessing the OCT images, we ran each of the 140 images through the ReLayNet algorithm and saved the *e*3, *bn*, and *d*3 intermediate convolutional layers as NumPy array data structures.

### SVM Classification

Support Vector Machines (SVMs) are machine learning models that can be used to analyze and classify data. To determine if the intermediate convolutional layers of the FEP and chronic schizophrenia patients differed from their age-matched controls, we utilized scikit-learn’s SVM module to build Support Vector Classification (SVC) models and see how accurately the models could classify an unseen OCT-derived data sample as being from a schizophrenia patient or control subject (Pedregosa et al., 2011). For comparison, we also built SVCs using the values provided by the Cirrus 5000 OCT machine. In total we analyzed 6 different data sets using SVCs: the features from the 1) *e*3, 2) *bn*, and 3) *d*3 convolutional layers, 4) all OCT machine-provided values, 5) the OCT machine-provided values that were analyzed in Lai et al., and 6) the OCT machine-provided values that were found to have significant differences between populations in Lai et al. (see Table 1).

**Table 1.**
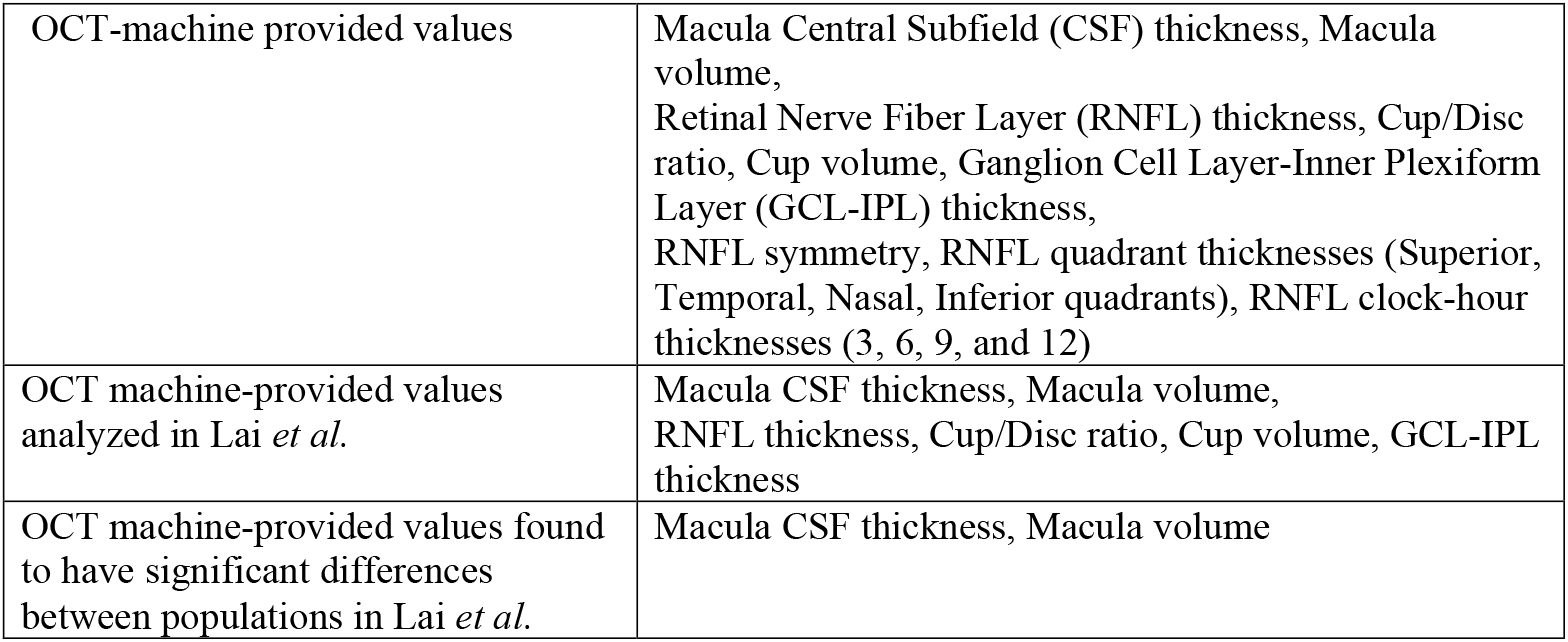
OCT machine-provided values used to create 3 different sets of SVCs.

Each of these data sets was partitioned such that SVCs were built to classify FEP patients vs. age-matched controls or chronically ill patients vs. age-matched controls. Given the small number of data samples available, we implemented a leave-one out cross-validation process. Specifically, we built SVCs by training the algorithm with all of the samples in a data subset, except one, and tested them by having the models label the unseen sample. For example, when using the *e*3 convolutional layers from the FEP and age-matched control OCT images, we created 68 classifiers because there were 14 FEP subjects and 20 controls, with 2 retina images each. The classifiers were all trained on 67 sets of features extracted from the training image set, and each was tested by labeling features from the remaining unseen image. This way, features extracted from every image were used to test a classifier that was trained on the remainder of the data.

### SVC Parameters and Data Preprocessing

One technique used to improve the performance of the SVCs was to adjust for the data imbalance of the training set. Because the groups being compared (i.e., FEP vs. control) did not have the same number of subjects (i.e., 14 vs. 20, each providing data from both eyes), an SVC could be biased to more likely label an unseen sample as belonging to whichever group had a larger sample size. To account for this, we used scikit’s `class_weights=balanced` parameter, which automatically balanced the weights of different groups so that an SVC was not influenced by their sizes. The equation for the weight of each group or class was as followed:

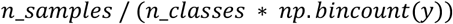

Another technique we tested was standardizing the data sets by removing the mean and adjusting the standard deviation of each variable to 1, using scikit-learn’s preprocessing `StandardScaler` function. Standardization was necessary for the SVCs created using OCT machine-provided values, because the low dimensionality of the data resulted in non-convergence, meaning differing results with repeated trials. Standardization was not necessary to achieve convergence on SVCs operating on features extracted from convolutional layers, so we created SVCs for these using both standardized and nonstandardized data sets.

### Data Analysis

Our labelling followed the traditional convention according to which “schizophrenia” samples belong to the positive class and “control” samples belong to the negative class. A true positive or true negative resulted when the test sample was correctly labeled as schizophrenia or control, respectively. For each set of classifiers (i.e., SVCs built using all OCT machine-provided data and labeling FEP vs. control), a confusion matrix was constructed outlining the number of true positives (TP), true negatives (TN), false positives (FP), and false negatives (FN) that resulted. We then calculated the accuracy, precision, recall, and *f*1 values for each confusion matrix, where each of these values is defined as followed:

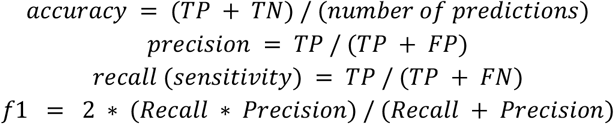

The accuracy and *f*1 values were used to compare the performance of the different sets of classifiers.

## Results

### SVCs trained on OCT Machine Values

Using OCT machine-provided values as features, SVCs that labeled chronic schizophrenia vs. age-matched control samples had good performance, based on their accuracy and *f*1 values, while SVCs labeling FEP vs. age-matched controls did not.

For the chronic schizophrenia vs. control classifiers, all of the SVCs resulted in both accuracies and *f*1 scores greater than 0.5, meaning that their performance was better than the expected result had the algorithms randomly assigned the two labels (see Table 2). The SVCs built and tested on only the macular thickness and cube volume values performed best (accuracy = 0.72, *f*1 = 0.70). The classifiers created using all OCT machine-provided values and only the provided values analyzed in Lai et al. performed comparably with equal accuracies (accuracy = 0.69) and *f*1 scores of 0.66 and 0.64, respectively.

**Table 2.** Confusion matrices and statistical analysis values for OCT machine data SVCs.

| FEP vs. Control |  |  |  |  | Chronic SZ vs. Control |  |  |  |  |
| --- | --- | --- | --- | --- | --- | --- | --- | --- | --- |
| All OCT Machine Data |  |  |  |  |  |  |  |  |  |
|  |  | Predicted Values |  |  |  |  | Predicted Values |  |  |
|  |  | Control | Patient | Total |  |  | Control | Patient | Total |
| Actual Values | Control | TN = 6 | FP = 14 | 20 | Actual Values | Control | TN = 14 | FP = 4 | 18 |
|  | Patient | FN =7 | TP = 7 | 14 |  | Patient | FN = 7 | TP = 11 | 18 |
|  | Total | 13 | 21 | 34 |  | Total | 21 | 15 | 36 |
|  | Accuracy: 0.38 | Precision: 0.33 | Recall: 0.5 | f1: 0.4 |  | Accuracy: 0.69 | Precision: 0.73 | Recall: 0.61 | f1: 0.66 |
| Analyzed OCT Machine Data |  |  |  |  |  |  |  |  |  |
|  |  | Predicted Values |  |  |  |  | Predicted Values |  |  |
|  |  | Control | Patient | Total |  |  | Control | Patient | Total |
| Actual Values | Control | TN = 6 | FP = 14 | 20 | Actual Values | Control | TN = 15 | FP = 3 | 18 |
|  | Patient | FN =5 | TP = 9 | 14 |  | Patient | FN = 8 | TP = 10 | 18 |
|  | Total | 11 | 23 | 34 |  | Total | 23 | 13 | 36 |
|  | Accuracy: 0.44 | Precision: 0.39 | Recall: 0.64 | f1: 0.48 |  | Accuracy: 0.69 | Precision: 0.76 | Recall: 0.55 | f1: 0.64 |
| Significant OCT Machine Data |  |  |  |  |  |  |  |  |  |
|  |  | Predicted Values |  |  |  |  | Predicted Values |  |  |
|  |  | Control | Patient | Total |  |  | Control | Patient | Total |
| Actual Values | Control | TN = 12 | FP = 8 | 20 | Actual Values | Control | TN = 14 | FP = 4 | 18 |
|  | Patient | FN =9 | TP = 5 | 14 |  | Patient | FN = 6 | TP = 12 | 18 |
|  | Total | 21 | 13 | 34 |  | Total | 20 | 16 | 36 |
|  | Accuracy: 0.5 | Precision: 0.38 | Recall: 0.35 | f1: 0.37 |  | Accuracy: 0.72 | Precision: 0.75 | Recall: 0.66 | f1: 0.70 |

For the FEP vs. control classifiers, there was no classifier that had both accuracy and *f*1 values greater than 0.5 or performed better than the others when looking at both fields (see Table 2). Classifiers that looked only at macular thickness and cube volume had the highest accuracy (accuracy = 0.5), and classifiers that only looked at values analyzed in Lai et al. had the highest *f*1 scores (*f*1 = 0.48).

### SVCs trained on Deep OCT Image Features

Comparing the overall results of SVCs looking at chronic schizophrenia patients to those looking at FEP schizophrenia patients, neither set of SVCs consistently outperformed the other.

For the chronic schizophrenia vs. control classifiers, the SVCs using features extracted from the *d*3 intermediate convolutional layers with non-standardized data had the best performance (accuracy = 0.62, *f*1 = 0.58, see Table 3). The SVCs relying on non-standardized data produced higher *f*1 scores than those using standardized data for all three convolutional layers. Also comparing only *f*1 scores, the *d*3 layer features had the best performance within the standardized and non-standardized categories.

**Table 3.**
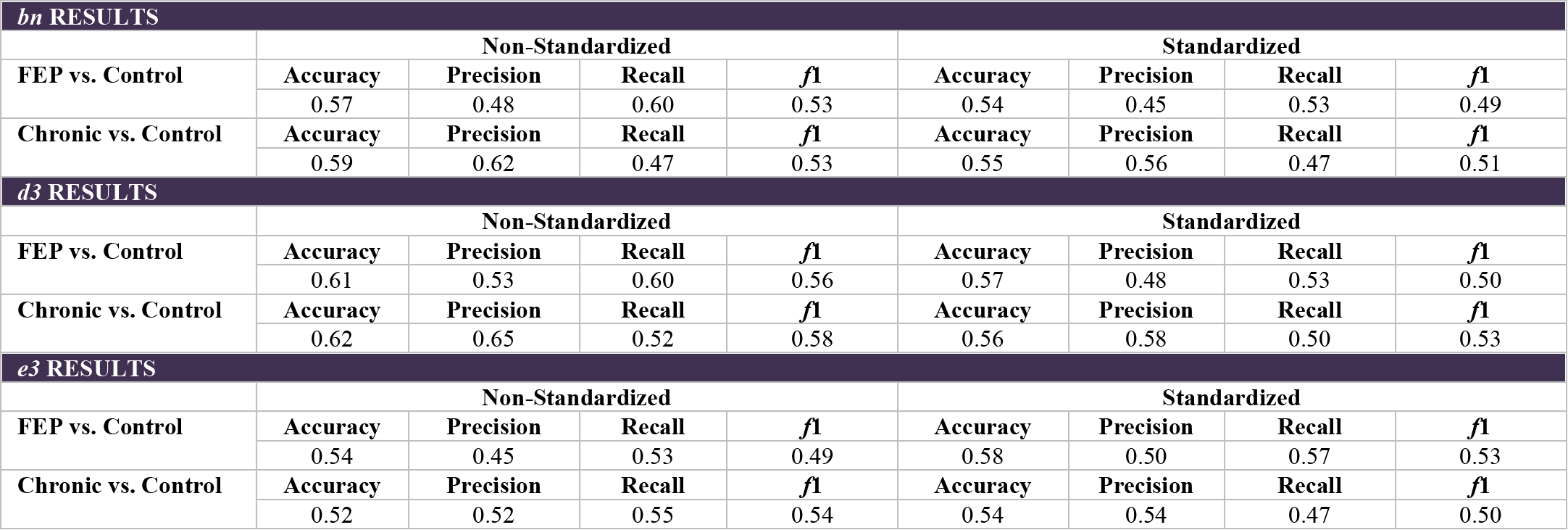
Statistical analysis values for OCT image-based SVCs.

For the FEP vs. control classifiers, the SVCs using features extracted from the *d*3 intermediate convolutional layers with non-standardized data once again had the best performance (accuracy = 0.61, *f*1 = 0.56, see Table 3). For features extracted from the *bn* and *d*3 layers, the non-standardized data outperformed the standardized data. However, for features extracted from the *e*3 convolutional layer, the standardized data performed better (standardized: *f*1 = 0.53, non-standardized: *f*1 = 0.49). Within the non-standardized data category, the *d*3 layer features still performed best, and within the standardized data category, the *e*3 layer features performed best (*f*1 = 0.53).

We hypothesize the reason why standardization of deep features does not have a significant impact on the experimental results is related to the fact that all feature values lie on the same feature space, and consequently, their magnitudes have comparable physical meaning. This is not the case for OCT machine values, which carry largely diverse physical meanings. Additionally, classifiers operating on features extracted from the *d*3 layers may outperform those leveraging earlier sets of deep features due to the fact that, the deeper the feature, the higher the level of abstraction associated with it. As mentioned, however, using features that are too close to the output layer may be counterproductive since their level of task specificity (in this case, segmentation) may prevent them from being effective at other tasks, such as the one this work focuses on.

Of note, for the FEP schizophrenia patient vs. control classifiers, the SVCs built using features extracted from OCT images consistently performed better than SVCs built using values provided by the OCT machine. In contrast, for the chronic schizophrenia vs. control classifiers, the SVCs built from OCT machine values consistently had higher *f*1 scores.

## Discussion

This study demonstrated the potential for ML analyses in research on retinal imaging in schizophrenia. The classifiers built using OCT machine metrics paralleled the findings in the Lai et al. study, with those differentiating chronic schizophrenia patients from controls performing with good accuracy while those differentiating FEP schizophrenia patients from controls had poor accuracy. This shows that relying on SVCs to analyze OCT data is equivalent to implementing parametric statistics. Building on this finding, we successfully created a set of classifiers trained on features extracted from OCT images that performed with accuracy above 0.5 for both the FEP and chronic schizophrenia populations, revealing the potential effectiveness of ML in analyzing OCT images and distinguishing differences between these groups. Although our accuracies of 0.61 and 0.62 yield room for improvement, what makes this a noteworthy finding is that the ML algorithm we used to extract OCT image data was not optimized for data from schizophrenia patients. Rather, due to the small data set, we used an algorithm based on diabetic macular edema patients that had been previously generated on a significantly larger, publicly available data set. In addition, the ReLayNet algorithm was optimized for a segmentation task, whereas we aimed to discriminate schizophrenia patients from controls. We hypothesize that accuracy would be significantly higher if a model trained for our goal and tested on a large sample of schizophrenia patients and controls could be developed.

This proof-of-concept study demonstrated that ML analyses of OCT data, especially those developed on a dataset of schizophrenia patients, could be useful in several respects. First, these may eventually be shown to be more sensitive than traditional OCT thickness and volume variables for identifying schizophrenia-related abnormalities. Second, once an optimal algorithm is identified, it can be used to discover those image features –and therefore, which anatomical features– are most associated with schizophrenia, thereby leading to advances in our understanding of pathophysiology. This could also potentially have implications for treatment and prevention of further decline in retinal and possibly brain health.

No study published to date on OCT findings in schizophrenia has had a sample size that is sufficiently large to be used for development of a schizophrenia-specific deep ML model. Because it is unlikely that any single study would be large enough, establishment of a network of clinical sites, collecting data on well-characterized patient samples using the same OCT equipment and software, and contributing to a centralized data registry, is a logical next step. In our view, the relative consistency of findings of retinal structural abnormalities in schizophrenia across studies, and the advantages of these data for revising our view of schizophrenia within the context of other neuropsychiatric disorders, warrants pursuing this approach.

## Data Availability

All data described in the manuscript are available from the authors upon request.

## VI. Acknowledgments

Medical Education Awards Program, University of Rochester School of Medicine and Dentistry

## Conflict of Interest Statement

Joseph, Lai, Silverstein, Ramchandran, & Bernal

Using Machine Learning to Classify Schizophrenia Based on Retinal Images

The authors report no conflicts of interest, or potential perceived conflicts of interest regarding this work.

